## Supplementary material for "COVID-19 mitigation measures in primary schools and association with infection and school staff wellbeing: an observational survey linked with routine data in Wales, UK": S5 Appendix

### Difficulty being heard/understood – having to talk louder

“*I often end up raising my voice more often*” (teaching assistant)

“*If I wear a mask whilst teaching I feel that it affects the children as they find it hard to understand what I am saying*” (support staff)

“*I feel that it hinders the communication between myself and the pupils. Pupils cannot always hear me and they are sometimes distracted by the appearance*” (teacher)

“*It makes you feel hot and you have to shout to be heard. It is a long day with masks and not very comfortable*” (teacher)

“*I absolutely hate wearing a mask or visor. It restricts me and it makes me feel poorly. You have to project your voice more as children don't hear you all the time*” (teacher)

### Difficulty understanding body language/facial expressions

“*Makes the children uncomfortable - hard to read facial expressions and to deliver lessons when wearing a mask*” (teacher)

*“Yes as an Early Years teacher I feel the children benefit enormously by seeing facial expression when teaching*” (teacher)

“*I find it extremely difficult to wear a mask/visor whilst teaching. They are young children and need to see facial expressions”* (teacher)

### Physical impacts of wearing including health and vision

“*My throat is constantly sore because you have to raise your voice to be heard over the shield. It also makes it difficult to hear*” (teacher)

“*It gets quite hot and hard to breathe sometimes, some children struggle with what is being said to them if the mask is on*” (teaching assistant)

“*It is awful breathing in my breath all day and this constantly happens, as teachers we are constantly talking. It is very difficult but I am getting used to it*” (teacher)

“*Yes, children can't hear you. You steam up. Outdoors visors get wet. It makes it very difficult to teach well*” (teacher)

“*I wear glasses so find both visor and masks hugely impact my vision due to condensation on my glasses with mask and poor visibility with visor*” (teaching assistant)

### Social/emotional impacts affecting relationships with pupils

“*I wear a mask in communal areas and a visor to teach the children. The children don't hear me as well and it makes teaching feel cold and uncaring*” (teacher)

“*Wearing a mask hides a smile and other facial expressions. I believe it's hard to read ones emotions with wearing a mask*” (teaching assistant)

### Challenges for pupils with additional learning needs and English as an additional language

*“Mask reduces audibility of voice and makes it impossible for children with speech and language difficulties to see how you articulate different sounds”* (teaching assistant)

*“Hard to support children with speech difficulties or hearing difficulties*” (headteacher)

### Impact on teaching phonics

“*It is difficult for the children to understand sometimes. It is very difficult teaching phonics, where mouth shape and lip position is important*” (teaching assistant)

“*Voice is more muffled, so it's difficult to teach phonics or read stories. I'm using my voice more to be heard*” (teacher)

*“When delivering phonics the children struggle when we are wearing masks as they can not see our mouth forming the words and sounds”* (role unknown)
