## Supplementary material for "COVID-19 mitigation measures in primary schools and association with infection and school staff wellbeing: an observational survey linked with routine data in Wales, UK": S4 Appendix

|  | Moderate/severe anxiety (GAD-7) (individual level: school staff) | | Moderate/severe depression (PHQ-9) (Individual level: school staff) | |
| --- | --- | --- | --- | --- |
| Self-reported measures from survey | **OR** | **95% CI** | **OR** | **95% CI** |
| *Face covering – MASK (reference no face covering)* | 1.02 | 0.53 to 1.97 | *1.86* | *0.97 to 3.57* |
| *Face covering – VISOR (reference no face covering)* | 2.28 | 0.82 to 6.35 | **4.19** | **1.40 to 12.53** |
| *Keep 2 metres from PUPILS – SOMETIMES (reference never/rarely)* | 0.66 | 0.33 to 1.35 | 1.13 | 0.53 to 2.39 |
| *Keep 2 metres from PUPILS – MOST THE TIME/ALWAYS (reference never/rarely)* | 2.05 | 0.75 to 5.60 | 1.74 | 0.67 to 4.55 |
| *Keep 2 metres from STAFF – SOMETIMES (reference never/rarely)* | 0.50 | 0.13 to 1.82 | 0.69 | 0.15 to 3.20 |
| *Keep 2 metres from STAFF – MOST THE TIME/ALWAYS (reference never/rarely)* | 0.72 | 0.22 to 2.31 | 0.59 | 0.14 to 2.55 |
| *Non-household contacts 1-metre: Up to 5 (reference 0)* | 0.92 | 0.40 to 2.12 | 1.73 | 0.71 to 4.18 |
| *Non-household contacts 1-metre: 6+ (reference 0)* | 1.37 | 0.59 to 3.19 | *2.04* | *0.95 to 4.38* |
| *Non-household contacts direct: Up to 5 (reference 0)* | 0.53 | 0.16 to 1.80 | 0.93 | 0.30 to 2.90 |
| *Non-household contacts direct: 6+ (reference 0)* | 1.83 | 0.59 to 5.63 | 0.95 | 0.40 to 2.38 |
| *Classes mix at play* | 1.06 | 0.51 to 2.23 | 0.90 | 0.39 to 2.08 |
| *School offers breakfast club* | 0.78 | 0.41 to 1.49 | 0.74 | 0.33 to 1.68 |
| *School offers extra-curricular clubs* | 1.28 | 0.51 to 3.19 | 0.83 | 0.22 to 3.15 |
| *Teach outdoors – SOMETIMES (reference never/hardly ever)* | 0.62 | 0.33 to 1.15 | 0.68 | 0.28 to 1.68 |
| *Teach outdoors – MOST OF THE TIME/ALWAYS (reference never/hardly ever)* | 0.65 | 0.24 to 1.74 | 1.19 | 0.32 to 4.37 |
