## Supplementary material for "COVID-19 mitigation measures in primary schools and association with infection and school staff wellbeing: an observational survey linked with routine data in Wales, UK": S2 Appendix

**HAPPEN School Staff Survey 2020/2021**

Items used within analyses denoted **bold**

You and your school

1. First name
2. Last name
3. Ethinicity
   i. *Asian*
   ii. *Black*
   iii. *White*
   iv. *Mixed*
   v. *Prefer not to say*
4. Date of birth
5. Postcode
6. House number
7. What school do you work in?
8. What type of school is this?
   i. *English medium primary school*
   ii. *Welsh medium primary school*
   iii. *Dual stream school*
   iv. *Pupil referral unit*
   v. *Voluntary aided school*
   vi. *Special needs school*
   vii. *Private school*
   viii. *Specialist teaching facility within primary school*
   ix. *Other*
9. What is your role?
   i. *Support staff*
   ii. *Teaching assistant*
   iii. *Headteacher – teaching*
   iv. *Headteacher – non teaching*
   v. *Teacher*
   vi. *Higher level teaching assistant*
   vii. *Supply teacher*
10. Do you work:
    i. *Full time*
    ii. *Part Time*
11. What year do you teach?
    i. *Reception*
    ii. *Year 1*
    iii. *Year 2*
    iv. *Year 3*
    v. *Year 4*
    vi. *Year 5*
    vii. *Year 6*
    viii. *Combination of year groups*
12. Are you currently:
    i. *In school*
    ii. *Off school due to coronavirus symptoms and awaiting results of test*
    iii. *Off school due to other reasons*
    iv. *Self isolating due to close contact with positive case in school*
    v. *Self isolating due to close contact with positive case out of school*
13. **In the past 7 days have you felt ill with cold symptoms?**
    i. ***Yes***
    ii. ***No***
14. How do you normally get to school?
    i. *By car*
    ii. *By bus*
    iii. *By bike*
    iv. *By walking*
    v. *By running*
    vi. *By taxi*
    vii. *By train*
15. Do you have children living with you?
    i. *No*
    ii. *Yes, I have children under the age of 12*
    iii. *Yes, I have children over the age of 12*
16. Have you been shielding or do you live with someone who was shielding?
    i. *No*
    ii. *Yes, I was shielding*
    iii. *Yes, I live with someone who was shielding*
17. **Over the last 2 weeks how often have you been bothered by any of the following problems? [options: Not at all, Several days, More than half the days, Nearly every day] (GAD-7)**
    i. ***Feeling nervous, anxious or on edge?***
    ii. ***Not being able to stop or control worrying?***
    iii. ***Worrying too much about different things?***
    iv. ***Trouble relaxing?***
    v. ***Being so restless that it is hard to sit still?***
    vi. ***Becoming easily annoyed or irritable?***
    vii. ***Feeling afraid as if something awful might happen?***
18. On a scale of 1 to 10, where 1 is “not at all” and 10 is “completely”, overall, how satisfied are you with your life nowadays? [options: scale 1-10]
19. Have you experienced any times of low mood recently?
    i. *Yes*
    ii. *No*
20. **Over the last two weeks, how often have you been bothered by any of the following problems? [Options: Not at all, Several days, More than half the days, Nearly everyday] (PHQ-9)**
    i. ***Little interest or pleasure in doing things?***
    ii. ***Feeling down, depressed, or hopeless?***
    iii. ***Trouble falling or staying asleep, or sleeping too much?***
    iv. ***Feeling tired or having little energy?***
    v. ***Poor appetite or overeating?***
    vi. ***Feeling bad about yourself - or that you are a failure or have let yourself or your family down?***
    vii. ***Trouble concentrating on things, such as reading the newspaper or watching television?***
    viii. ***Moving or speaking so slowly that other people could have noticed? Or the opposite - being so fidgety or restless that you have been moving around a lot more than usual?***
    ix. ***Thoughts that you would be better off dead, or of hurting yourself in some way?***
21. What approach has your school taken to returning? e.g. is there any change in number of contacts between children and/or teachers since before COVID-19? How often do children and staff need to gel/wash hands? How are breaktimes managed for both pupils and staff? [Open text]
22. **Is your school offering breakfast club?**
    i. ***Yes***
    ii. ***No***
23. **Is your school offering after school club/s?**
    i. ***Yes***
    ii. ***No***
24. **Since your return to school, how much are you currently delivering teaching outdoors? (outdoor learning)**
    i. ***Most of the time***
    ii. ***Some of the time***
    iii. ***Hardly ever***
    iv. ***Never***
25. And is this more or less than before lockdown?
    i. *Teaching outdoors more*
    ii. *Teaching outdoors less*
    iii. *Teaching outdoors the same*
26. How much play time are pupils currently having?
    i. *More than before lockdown*
    ii. *The same as before lockdown*
    iii. *Less than before lockdown*
27. **Are different classes in your school mixing during playtimes?**
    i. ***Yes, in the hall***
    ii. ***Yes, outdoors in a field or large outdoor space***
    iii. ***No, there is no mixing of classes***
28. **Do you wear a mask in school?**
    i. ***No***
    ii. ***I wear a mask in communal areas but not in the classroom***
    iii. ***I wear a mask when in the classroom teaching or supporting***
    iv. ***I wear a visor in communal areas but not in the classroom***
    v. ***I wear a visor when in the classroom teaching or supporting***
29. **If you do wear a mask/visor do you feel it affects your teaching and in what way?**
30. How are pick ups and drop offs managed at your school? (tick all that apply)
    i. *The different years have staggered pick up and drop off times and pupils cannot enter the classroom before their allocated time*
    ii. *The different years have staggered pick up and drop off times but siblings can go into their classroom and wait if earlier drop off in their family*
    iii. *Parents are asked to wait behind a marked off area*
    iv. *Children are not allowed to enter the school until just before classes start*
    v. *There is no change compared to before COVID-19*
31. Do you feel current guidelines are affecting your teaching and pupil learning? If so, how? [open text]
32. Do you feel current guidelines are affecting the wellbeing of children? If so, how? [open text]
33. **How often are you able to keep 2 metres from children you are teaching or supporting?**
    i. ***Never***
    ii. ***Rarely***
    iii. ***Some of the time***
    iv. ***Most of the time***
    v. ***Always***
34. **How often are you able to keep 2 metres away from other members of staff during the teaching day?**
    i. ***Never***
    ii. ***Rarely***
    iii. ***Some of the time***
    iv. ***Most of the time***
    v. ***Always***
35. **How many people (who are not part of your household) did you talk to yesterday (e.g. were within 1 metre and exchanged a few words but did not touch)? [open text]**
36. **How many people did you have direct physical contact with yesterday (e.g. hugged, touched, kissed) who were not part of your household? [open text]**
37. Do you feel you were able to answer the last two questions above accurately?
    i. *Yes I can remember very well who I had contact with yesterday*
    ii. *No not really really, I am guessing*
    iii. *I honestly cant remember, I would say these answers are not very accurate*
38. What support is there for the health and wellbeing of staff in your school? [open text]
39. What would you change to improve the health and wellbeing of staff in your school? [open text]
40. What support is there for the health and wellbeing of pupils in your school? [open text]
41. What would you change to improve the health and wellbeing of pupils in your school? [open text]
