## Supplementary material for "COVID-19 mitigation measures in primary schools and association with infection and school staff wellbeing: an observational survey linked with routine data in Wales, UK": S3 Appendix

| Exposures | Survey item | Survey responses categories | Coding for analyses |
| --- | --- | --- | --- |
| Keep two metres from PUPILS | *33. How often are you able to keep 2 metres from children you are teaching or supporting?* | *i) Never*  *ii) Rarely*  *iii) Some of the time*  *iv) Most of the time*  *v) Always* | Ordinal:   - *Never/rarely (i, ii)* - *Some of thetime (iii)* - *Most of the time/always (iv, v)* |
| Keep two metres from STAFF | *34. How often are you able to keep 2 metres away from other members of staff during the teaching day?* | *i) Never*  *ii) Rarely*  *iii) Some of the time*  *iv) Most of the time*  *v) Always* | Ordinal:   - *Never/rarely (i, ii)* - *Some of the time (iii)* - *Most of the time/always (iv, v)* |
| Wear face covering (mask, visor) | *28. Do you wear a mask in school?* | i) No  ii) I wear a mask in communal areas but not in the classroom  iii) I wear a mask when in the classroom teaching or supporting  iv) I wear a visor in communal areas but not in the classroom  v) I wear a visor when in the classroom teaching or supporting | Nominal:   - *No (i)* - *Yes – mask (ii, iii)* - *Yes – visor (iv, v)* |
| Number of non-household contacts 1-metre | *35. How many people (who are not part of your household) did you talk to yesterday (e.g. were within 1 metre and exchanged a few words but did not touch)?* | Continuous number | Ordinal:   - *0* - *1-5* - *6+* |
| Number of non-household contacts direct | *36. How many people did you have direct physical contact with yesterday (e.g. hugged, touched, kissed) who were not part of your household?* | Continuous number | Ordinal:   - *0* - *1-5* - *6+* |
| Different classes mixing at play | *27.* *Are different classes in your school mixing during playtimes?* | i) Yes, in the hall  ii Yes, outdoors in a field or large outdoor area  iii) No, there is no mixing of classes | Binary:   - *Yes (i, ii)* - *No (iii)* |
| School offers breakfast club | *22. Is your school offering breakfast club?* | i) Yes  ii) No | Binary:   - *Yes (i)* - *No (ii)* |
| School offers extra-curricular clubs | *23. Is your school offering after school club/s* | i) Yes  ii) No | Binary:   - *Yes (i)* - *No (ii)* |
| Teach outdoors | *26. Since your return to school, how much are you currently delivering teaching outdoors? (outdoor learning)* | i) Never  ii) Hardly ever  iii) Some of the time  iv) Most of the time | Ordinal:   - *Never/hardly ever (i, ii)* - *Some of the time (iii)* - *Most of the time (iv)* |
